# Estimating the dietary and health impact of implementing mandatory front-of-package nutrient disclosures in the US: a policy scenario modeling analysis

**DOI:** 10.1101/2024.10.12.24315377

**Authors:** Nadia Flexner, Daniel Zaltz, Eva Greenthal, Aviva A. Musicus, Mavra Ahmed, Mary R. L’Abbe

## Abstract

**Background:** Recognized as a cost-effective policy to promote healthier diets, mandatory front-of-package labeling (FOPL) identifying foods high in sodium, sugar, and saturated fat has been adopted and implemented in ten countries, and is currently under consideration in several others including the US. However, its potential impact on dietary intake and health have not yet been estimated in the US context.

**Objectives:** To estimate (1) the potential dietary impact of implementing mandatory nutrient-specific FOPL among US adults; and (2) the number of diet related non-communicable disease (NCD) deaths that could be averted or delayed due to estimated dietary changes.

**Methods:** Baseline and counterfactual dietary intakes of sodium, sugars, saturated fats, and calories were estimated among US adults (n=7,572) using both available days of 24h recall data from the 2017-2020 National Health and Nutrition Examination Survey (NHANES). The National Cancer Institute method was used to estimate usual intakes and distributions, adjusting for age, sex, misreporting status, weekend/weekday, and sequence of recall. To estimate counterfactual dietary intakes, we modeled reductions observed in experimental and observational studies that examined changes in sodium, sugars, saturated fat and calorie content of food and beverage purchases due to nutrient-specific ‘high in’ FOPL. This study used the Preventable Risk Integrated ModEl (PRIME) to estimate potential health impact.

**Results:** Estimated mean dietary reductions of 156 mg and 259 mg/day of sodium, 10.1 g and 7.2 g/day of sugars, 1.08 g and 4.49 g/day of saturated fats, and 38 kcal and 57 kcal/day of calories were observed under the two policy scenarios tested. Between 96,926 (95% UI 89,011-105,284) and 137,261 (95% UI 125,534-148,719) diet related NCD deaths, primarily from cardiovascular diseases (74%), could potentially be averted or delayed by implementing mandatory nutrient-specific FOPL in the US. Overall, more lives would be saved in males than females.

**Conclusions:** Findings suggest that implementing mandatory nutrient-specific FOPL in the US could significantly reduce sodium and total sugar intakes among US adults, resulting in a substantial number of NCD related deaths that could be averted or delayed. Our results can inform current food policy developments in the US regarding adoption and implementation of FOPL regulations.

## Introduction

Poor dietary health characterized by an excess intake of nutrients-of-concern (i.e., sugar, sodium, and saturated fat), as well as underconsumption of whole grains, fruits, and vegetables, is a leading cause of global morbidity and mortality[1]. A major contributor to poor dietary intake across populations is the increasing availability and consumption of highly processed foods that are energy-dense and high in nutrients-of-concern, underscoring the importance of broad public health policies targeting both consumer and corporate behavior[2]. Front-of-package nutrition labeling (FOPL) on packaged foods and beverages is increasingly used as one such policy intervention to encourage healthier choices while simultaneously influencing food and beverage manufacturers to formulate healthier products[3]. FOPL provides consumers with clear and easy-to-understand nutritional information in addition to the nutrition information panels on the back or side of packages. For instance, some countries have adopted voluntary FOPL that summarize the healthfulness of products based on nutrients-of-concern and nutrients and ingredients to encourage (e.g., Nutriscore in Europe and Health Star Ratings in Australia and New Zealand). In contrast, other countries have adopted mandatory nutrient-specific FOPL policies, requiring labels on packaged, processed foods that are high in nutrients-of-concern, such as saturated fat, sugars, and sodium (e.g., ‘high in/excess’ labels in the Americas region)[4, 5].

Evidence from experimental studies has shown promising findings from implementing nutrient-specific ‘high in’ FOPL[6, 7], which has been confirmed by early evaluations of this policy in Chile[8–13], the first country to adopt and implement a nutrient-specific ‘high in’ FOPL in 2016[14]. The comprehensive Chilean Law also includes marketing restrictions and a ban on sales and promotion in schools for products that exceed established thresholds for calories and nutrients-of-concern[14]. Initial evaluations of the Chilean law have shown effectiveness in changing consumer purchasing behaviors[8, 9, 13], promoting industry-driven food reformulation[10, 11], decreasing unhealthy food advertising[15], and improving diets among children and adolescents[12]. For instance, after FOPL was implemented in Chile, the proportion of foods and beverages requiring at least one ‘high in’ FOPL decreased from 51% to 44%[11]. Furthermore, the policy led to a reduction in the purchase of ‘high in’ products, resulting in declines in purchased calories and nutrients-of-concern[9, 13].

Designing FOPL interventions requires developing evidence-based nutrient profiling models (NPMs) to categorize foods and beverages by their nutrient levels and/or ingredients, and applying established nutrient thresholds to determine which products would display a FOPL. Evidence suggests that manufacturers are motivated to reformulate products in response to these nutrient thresholds (i.e., to avoid ‘high in’ FOPL)[3, 10, 11]; therefore, a transparent process is needed when developing NPMs. A recent systematic review of NPMs developed for nutrition-related policies and regulations found that, in recent years, the proportion of NPMs that include nutrients to encourage has decreased substantially[16], which aligns with public health recommendations and best practices for developing NPMs[17]. An NPM is a key component of an effective FOPL policy. It also can be used to inform other healthy eating policies—as has been done in other countries—by identifying packaged foods and beverages that may be taxed or restricted from being advertised and promoted to children, available in schools, or included in federal food procurement[3, 16].

Introducing a government-led FOPL intervention in the US may be an important public health intervention to improve the population’s diet and health. The US Food and Drug Administration (FDA) committed to developing a FOPL system in 2022[18], and is currently conducting consumer research and soliciting stakeholder feedback to further this objective[19]. However, little is known about what potential diet and health impacts mandatory nutrient-specific FOPL may have on the US population. Therefore, the purpose of this study was to estimate the potential dietary impact of implementing mandatory nutrient-specific FOPL among US adults, and to estimate the number of diet related non-communicable disease (NCD) deaths that could be averted or delayed due to these dietary changes.

## Methods

### Overview

This is an NCD policy scenario modeling study which estimates potential changes in dietary intake and disease morbidity and mortality in response to FOPL in the US. Broadly, we first estimate usual dietary intake using nationally-representative intake data; then, we calculate expected changes in this usual intake in response to FOPL, using previously-published data. Finally, we use these expected changes in usual dietary intake to estimate the potential number of diet related NCD deaths that could be averted or delayed using the Preventable Risk Integrated ModEl (PRIME)[20].

### Dietary intake, demographic, and anthropometric data

We used 24-hour recalls (24HR), demographics, and anthropometrics from the most recent publicly-available National Health and Nutrition Examination Survey (NHANES) data collected between 2017-2020 (pre-pandemic)[21]. NHANES is a complex, multistage, clustered survey conducted biannually, but the most recent pre-pandemic cycle (2019-2020) was truncated and therefore combined with the prior cycle (2017-2018)[21]. The survey is designed to assess the health and nutritional status of non-institutionalized children and adults in the US. For this study, we used dietary intake data, demographics (age, sex, pregnancy, breastfeeding), and anthropometrics (height and body mass index, BMI) for adults only (≥19 y). The NHANES dietary intake data are collected through two 24HR, the first conducted in person within Mobile Examination Centers and the second, for a subsample, via telephone 3-10 days later. For this analysis we used both available 24HR.

### Study sample

We excluded individuals who reported daily calories below 500 or above 8000 (n=221)[22], those younger than 19 years (n=4,541), those pregnant or breastfeeding (n=77), and those with body mass index (BMI) less than 18.5 (n=223). The final analytic sample size was comprised of 7,572 individuals, of whom 6,556 (86.3%) had two 24-hour recalls (**Figure 1**).

**Figure 1.**
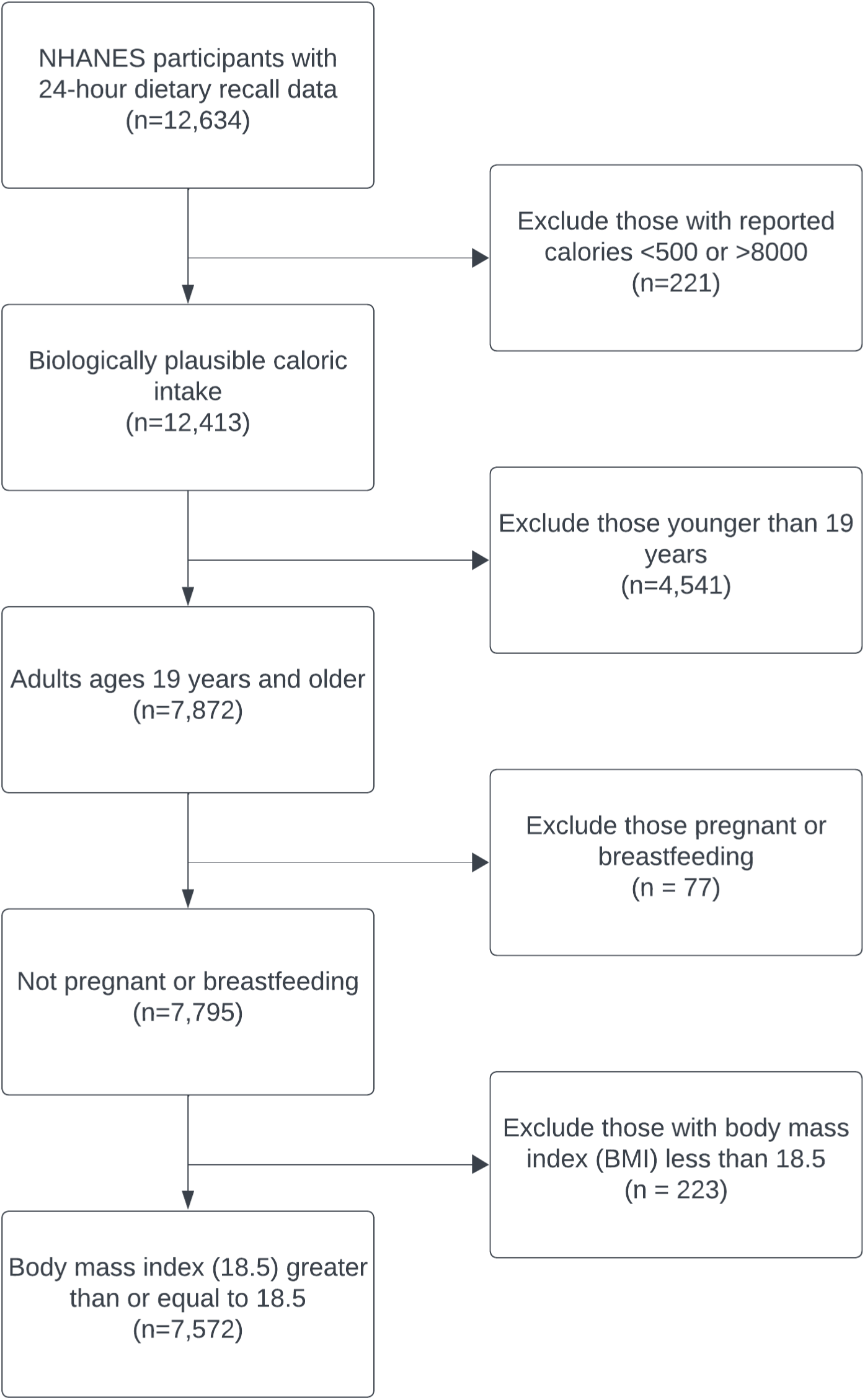
Study selection criteria, National Health and Nutrition Examination Survey, 2017 – 2020[21].

### Nutrient intakes and counterfactual scenarios

We focused on nutrients that the Dietary Guidelines for Americans recommend to limit in the diet[23], are used in FOPL from other countries [4, 16], and are highlighted in the latest FOPL schemes drafted by FDA[19, 24]. We assessed usual intakes of total energy (kcal/day), sodium (mg/day), total sugars (g/day), added sugars (g/day), saturated fats (g/day), and the proportion of total energy from saturated fats (%kcal/day). We defined the baseline scenario as the observed usual intake of each of the aforementioned nutrients within our study sample. Then, we obtained estimates of the effect of FOPL interventions on dietary intake based on current evidence to define two plausible population-level responses to mandatory FOPL in the United States(**Table 1**)[6, 9].

**Table 1.** Counterfactual scenarios: proportional change of calorie and critical nutrients content of food and beverage purchases in the presence of nutrient-specific ‘high in’ FOPL.

| Scenarios | Source | Tested foods | Main analysis |  |  |
| --- | --- | --- | --- | --- | --- |
| | | | Δsodium | Δsugars | Δsat fat |
| Scenario 1 (S1) | Based on early FOPL evaluations from Chile [9] | Food & beverages overall | -4.7% | -10.2% | -3.9% |
| Scenario 2 (S2) | Based on results from a systematic review and meta-analysis [6] | Food & beverages overall | -7.8% | -7.3% | -16.3% |

S1 and S2 analyze the potential dietary impact as a result of changes in consumer food purchases in the presence of mandatory nutrient-specific ‘high in’ FOPL.

#### Scenario 1: based on post-policy evaluation of mandatory ‘high in’ FOPL in Chile

Scenario 1 (S1) was informed by a post-policy evaluation of mandatory nutrient-specific ‘hig in’ FOPL in Chile. Early evaluations conducted a year after implementation examined changes in the calorie, sugar, sodium, and saturated fat content of food and beverage purchases[9].

Findings from this evaluation showed statistically significant declines in overall household purchases of saturated fat (−3.9%), sodium (−4.7%), and sugars (−10.2%) in the first 18 months after the policy was implemented[9]. For our counterfactual scenario, we systematically applied these overall reductions in purchases of saturated fats, sodium and sugars to the consumption of all foods and beverages reported in NHANES (2017-2020), except for dietary supplements and alcoholic beverages, as these are not included in FOPL regulations in countries that have implemented FOPL to date. We assumed that changes in the US would be of similar magnitude to the observed decreases in purchases of products ‘high in’ nutrients-of-concern and increases in purchases of products displaying no ‘high in’ FOPL in Chile[9].

#### Scenario 2: based on a meta-analysis on the impact of nutrient warning FOPL

Scenario 2 (S2) was informed by findings from a meta-analysis of randomized controlled trials and quasi-experimental studies that investigated overall changes in the calorie, sugar, sodium, and saturated fat content of food and beverage purchases in the presence of nutrient warning FOPL [6]. Specifically, we used the results comparing nutrient warning label vs. control/no label or Nutrition Facts table. Overall changes were applied for saturated fat (−16.3%), sodium (−7.8%), and sugars (−7.3%) to all foods and beverages reported in NHANES (2017-2020), except for dietary supplements and alcoholic beverages.

We estimated usual intake (described below) across each baseline and counterfactual scenario for the total study sample and provide in the supplemental material these results separately by Dietary Reference Intakes (DRI) age-sex groups[25]. Meaningful differences between observed intakes (baseline) and projected dietary intakes (counterfactual) were evaluated using non-overlapping 95th percentile confidence intervals of the means, similar to previous studies on dietary patterns [26, 27]. In both scenarios examined in this study, changes in calorie intake were determined by calculating the calories derived from changes in sugar and saturated fat content (i.e., counterfactual kcals = baseline kcals - ((Δsugars (g) * 4 kcal) + (Δsaturated fat (g) * 9 kcal))), aiming to maintain a conservative approach in our estimations, as reported changes in calories were higher in both studies informing S1 and S2.

### Sensitivity analysis

For both scenarios tested in this study, we considered changes in total sugars intake; however, given that added sugars are considered one of the nutrients-of-concern in the Dietary Guidelines for Americans, we also applied potential changes to only added sugars. We estimated potential dietary and health gains based on intakes of added sugars from NHANES 2017-2020 for baseline and counterfactual scenarios (included in the supplemental materials). However, it is worth noting that the potential FOPL intervention effect for both main scenarios in this analysis was based on changes in total sugars intake; therefore, any reduction specifically in added sugars is accounted for in the main scenarios.

### Health impact modeling

This study used PRIME to estimate the potential health impact (i.e., potential deaths averted or delayed) of estimated dietary intake changes (intervention effect)[20]. PRIME is a comparative risk assessment model that examines the potential impact of population-level changes in the distribution of behavioral risk factors on NCDs related mortality[20]. The model allows for the investigation of changes in behavioral dietary risk factors (i.e., energy, fruits & vegetables, fiber, salt, total fat, saturated fat, unsaturated fat and cholesterol), which can impact various health outcomes, including CVDs, cancers, diabetes, kidney disease, and liver disease (included in this study). PRIME does not include sugars as a dietary risk factor; rather, their potential effects on health outcomes are mediated through calorie reduction. Developed by researchers at the University of Oxford, the model has been widely used in different country contexts and has been endorsed by the WHO Regional Office for Europe[28]. Specifically, PRIME answers the question, “How many deaths would have occurred in the baseline year if the distribution of risk factors had been different?”[28]. Like other cross-sectional NCD scenario models, PRIME does not account for the effect of time lag between the exposure and disease outcome; therefore, it remains uncertain how long after the change in risk factor exposure the estimated health benefits would occur. The strengths and limitations of PRIME have been previously described in detail[20].

Model inputs included age- and sex-specific data from the US, including: 1) the number of people in the population (2019)[29]; 2) the number of diet related NCD deaths relevant to the study (2019)[30]; 3) baseline calorie and nutrients-of-concern dietary intake (2017-2020)[21]; 4) counterfactual calorie and nutrients-of-concern dietary intake (estimated intervention effect). Data on diet related NCD mortality were based on the WHO International Classification of Diseases 10 (ICD 10)[31].

### Statistical analysis

We described all continuous and categorical variables using measurements of central tendencies and frequencies. Then, we applied the estimated changes in each selected nutrient for both counterfactual scenarios to each individual’s observed intake in the data; each individual therefore had three measurements of nutrient intake: (1) their observed (“baseline”) intake from 24-hour recalls conducted during NHANES data collection, (2) their change in the respective nutrient based on estimates from scenario 1, and (3) their change in the respective nutrient based on estimates from scenario 2.

For each baseline or counterfactual scenario, we estimated usual intake using the National Cancer Institute method for ubiquitously consumed nutrients[32]. This method uses two-part modeling on with person-level random effects and variance estimations with post-stratified balanced repeated replication weights. Details of this method are available elsewhere[33]. We estimated these usual intakes for our total sample and within age and sex strata defined by DRI age-sex groupings[25], and report means, standard errors, and 95% confidence intervals. We conducted all usual intake analyses using SAS software 9.4 (SAS Institute Inc., Cary, NC, USA) with publicly available macros provided by the National Cancer Institute[32].

Then, using these estimates of usual intake at baseline and under the two counterfactual scenarios, we estimated the potential number of diet related NCD deaths that could be averted or delayed in the US. Therefore, for the simulation modeling aspect of this study, once all necessary data was inputted into PRIME, the model estimated changes in the number of diet related NCD deaths between the baseline and each counterfactual scenario tested. To gauge the uncertainty surrounding the outcomes, 95% uncertainty intervals (UIs) were estimated using Monte Carlo analysis within PRIME, with 10,000 iterations performed. This allowed for the epidemiological parameters to vary randomly based on the distributions established in the model from the literature[20]. PRIME then provided estimates of the diet related NCD deaths that could have been prevented or postponed for each of the tested counterfactual scenarios, both overall and disaggregated by sex and specific diseases under examination.

## Results

### Study Sample

Study sample characteristics are shown in **Supplementary Table S1**. Briefly, a total of 7,572 (≥19 y) NHANES (2017-2020) respondents were included in this analysis, 51% being females, and 42% having at least a high school diploma or equivalent. Furthermore, 25% of respondents had normal BMI, 32% overweight, and 44% obesity, according to the WHO definition[34].

### Baseline scenario: observed usual intake

The mean (SE) usual total energy intake per day among adults in the US ages 19 years and older was 2,077 (17) kcal (**Table 2**). Among these study participants, the mean (SE) daily intake of sodium was 3,349 (27) mg; total sugars 101.1 (1.5) g; saturated fats 27.6 (0.3) g; and percent of total energy from saturated fats 11.8% (0.1). Calorie and nutrient intakes for US adults overall and by DRI age-sex group are detailed in **Supplementary Table S2**.

**Table 2.**
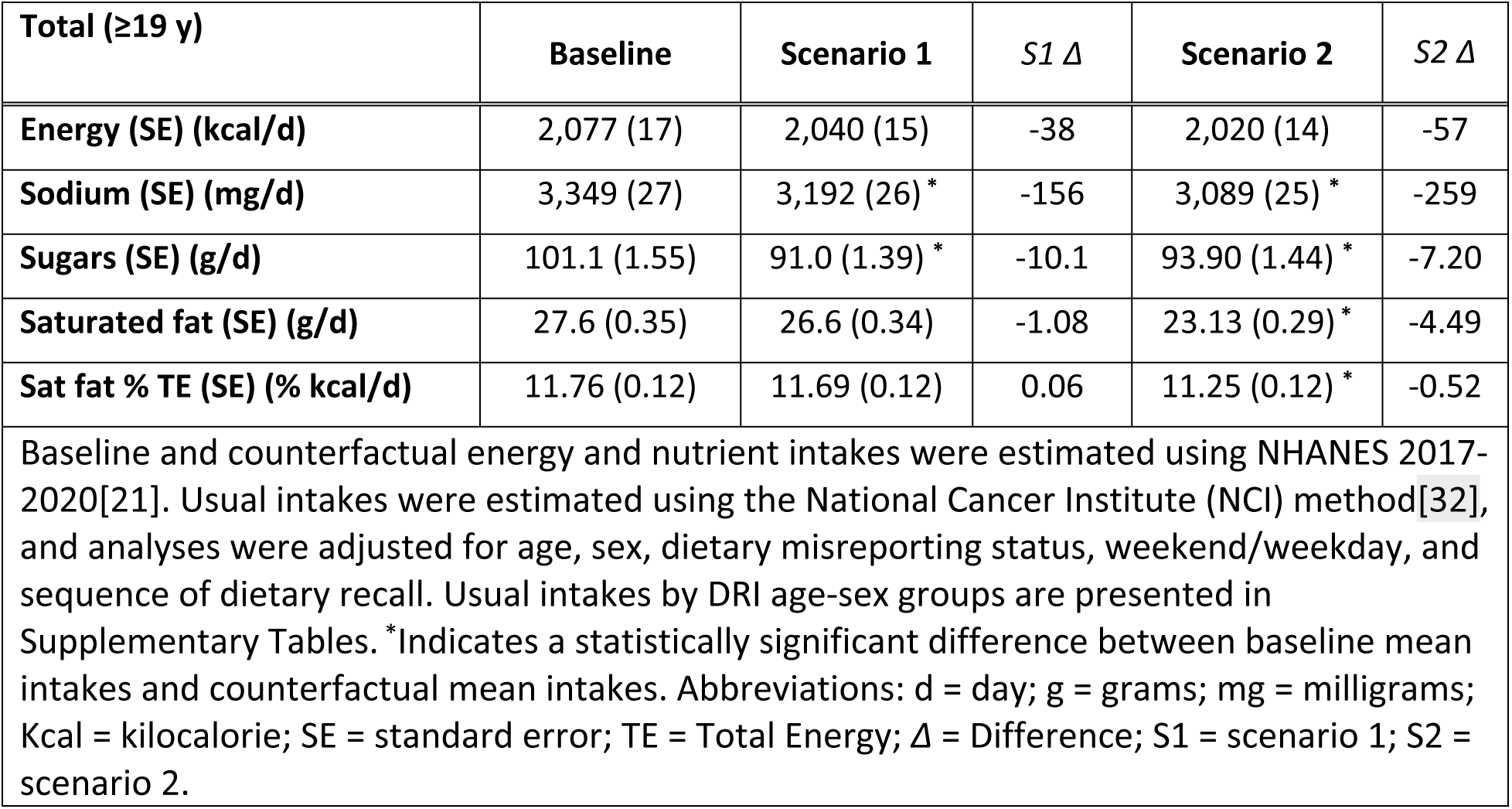
US adults’ (≥19 y) usual mean energy and nutrient intakes compared with estimated ‘counterfactual scenario’ mean intakes (n=7,572)

### Counterfactual scenarios: projected usual intake and health impact

#### Scenario 1: based on post-policy evaluation of mandatory ‘high in’ FOPL in Chile

Under the first counterfactual scenario (S1) based on results from the 18-month post-policy evaluation of mandatory FOPL in Chile[9], we projected a mean (SE) usual total energy intake of 2,040 (15) kcal per day. This represents a mean reduction of 38kcal/ day compared to the observed (baseline) scenario, considering only calorie contribution from changes observed in sugar and saturated fat. However, this reduction was not statistically significant. Additionally, we observed absolute mean dietary reductions of 156 mg/day sodium (−4.7%), 10.1 g/day sugars (−9.9%), and 1.1 g/day saturated fats (−3.9%). The reductions in sodium and sugar intakes were statistically significant (**Table 2**). Stratified results by DRI age-sex group were also estimated and are presented in **Supplementary Table S2**.

Estimated dietary intakes changes could avert or delay 96,926 (95% UI 89,011, 105,284) diet related NCD deaths in the US. Overall, more lives would be saved in males than females. Approximately 58% of averted or delayed deaths were estimated in males (56,536 [95% UI 51,524, 61,380]) and 42% in females (40,285 [95% UI 36,593, 44,050]) (**Table 3**).

**Table 3.**
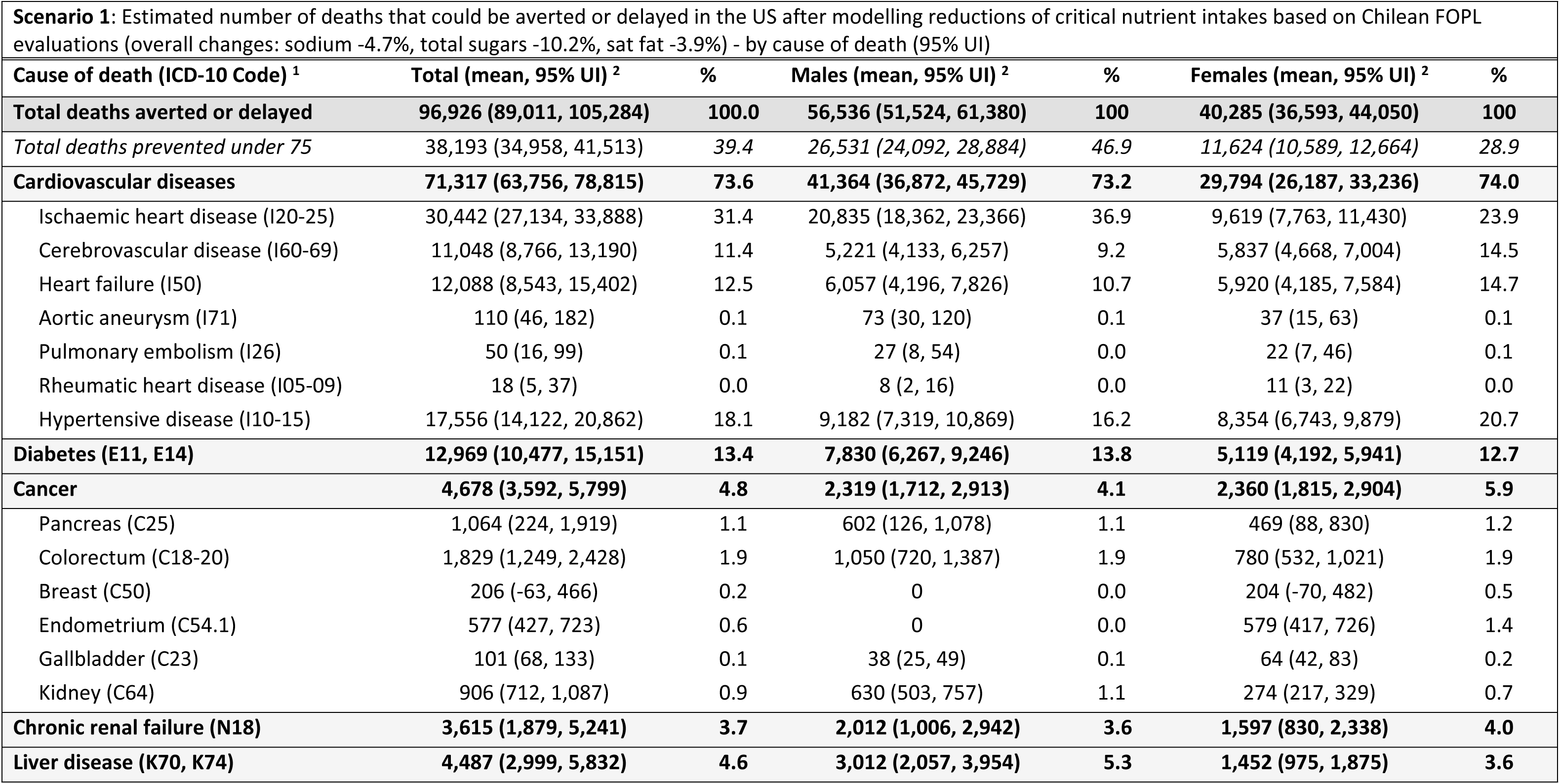
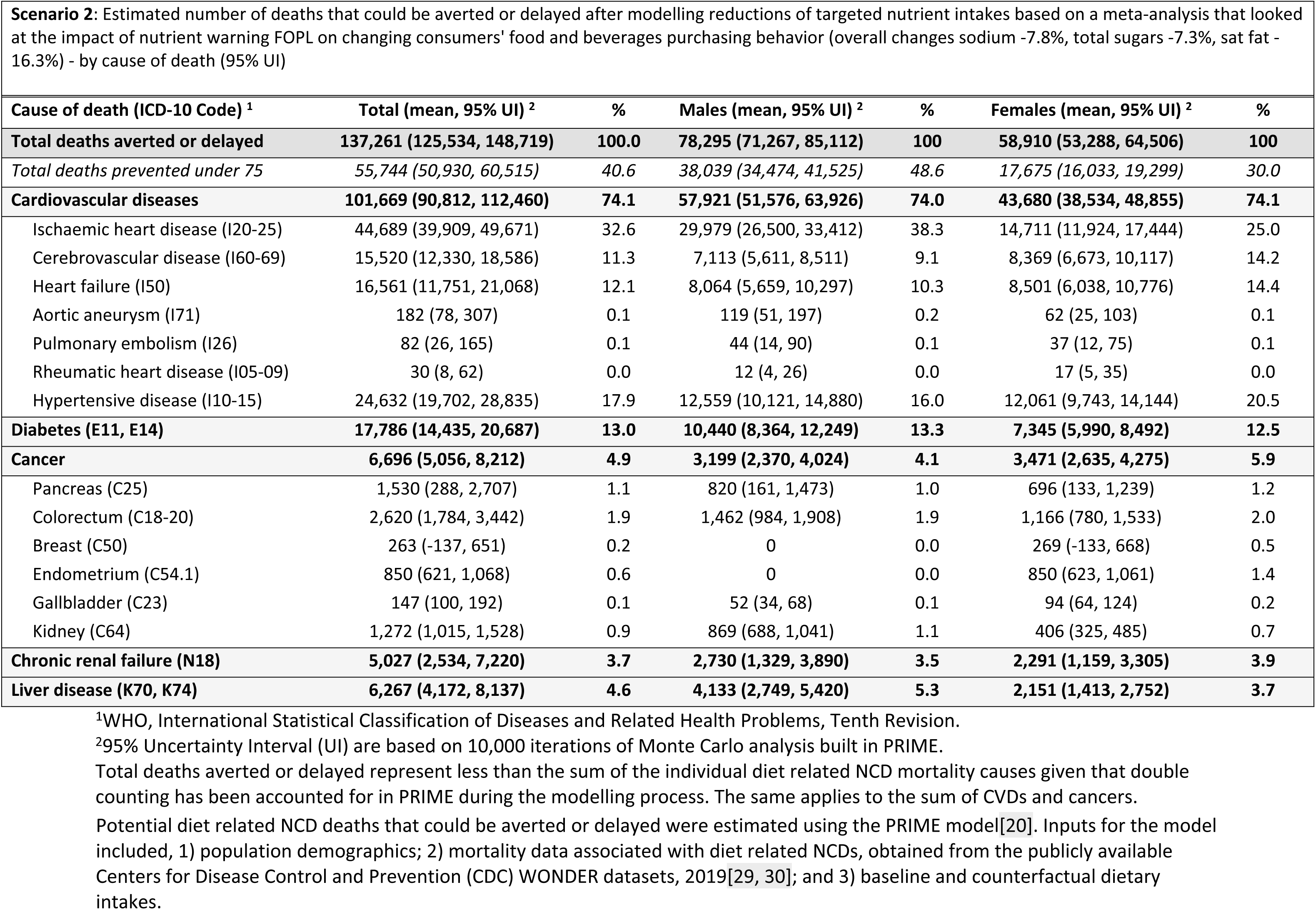
Diet related NCD deaths that could be averted or delayed in the US due to changes in calorie and critical nutrients content of food and beverage purchases in the presence of nutrient-specific ‘high in’ FOPL.

| **Scenario 1:** Estimated number of deaths that could be averted or delayed in the US after modelling reductions of critical nutrient intakes based on Chilean FOPL evaluations (overall changes: sodium -4.7%, total sugars -10.2%, sat fat -3.9%) - by cause of death (95% UI) |  |  |  |  |  |  |
| --- | --- | --- | --- | --- | --- | --- |
| **Cause of death (ICD-10 Code) <sup>1</sup>** | **Total (mean, 95% UI) <sup>2</sup>** | **%** | **Males (mean, 95% UI) <sup>2</sup>** | **%** | **Females (mean, 95% UI) <sup>2</sup>** | **%** |
| **Total deaths averted or delayed** | **96,926 (89,011, 105,284)** | **100.0** | **56,536 (51,524, 61,380)** | **100** | **40,285 (36,593, 44,050)** | **100** |
| *Total deaths prevented under 75* | 38,193 (34,958, 41,513) | 39.4 | 26,531 (24,092, 28,884) | 46.9 | 11,624 (10,589, 12,664) | 28.9 |
| **Cardiovascular diseases** | **71,317 (63,756, 78,815)** | **73.6** | **41,364 (36,872, 45,729)** | **73.2** | **29,794 (26,187, 33,236)** | **74.0** |
| Ischaemic heart disease (I20-25) | 30,442 (27,134, 33,888) | 31.4 | 20,835 (18,362, 23,366) | 36.9 | 9,619 (7,763, 11,430) | 23.9 |
| Cerebrovascular disease (I60-69) | 11,048 (8,766, 13,190) | 11.4 | 5,221 (4,133, 6,257) | 9.2 | 5,837 (4,668, 7,004) | 14.5 |
| Heart failure (I50) | 12,088 (8,543, 15,402) | 12.5 | 6,057 (4,196, 7,826) | 10.7 | 5,920 (4,185, 7,584) | 14.7 |
| Aortic aneurysm (I71) | 110 (46, 182) | 0.1 | 73 (30, 120) | 0.1 | 37 (15, 63) | 0.1 |
| Pulmonary embolism (I26) | 50 (16, 99) | 0.1 | 27 (8, 54) | 0.0 | 22 (7, 46) | 0.1 |
| Rheumatic heart disease (I05-09) | 18 (5, 37) | 0.0 | 8 (2, 16) | 0.0 | 11 (3, 22) | 0.0 |
| Hypertensive disease (I10-15) | 17,556 (14,122, 20,862) | 18.1 | 9,182 (7,319, 10,869) | 16.2 | 8,354 (6,743, 9,879) | 20.7 |
| **Diabetes (E11, E14)** | **12,969 (10,477, 15,151)** | **13.4** | **7,830 (6,267, 9,246)** | **13.8** | **5,119 (4,192, 5,941)** | **12.7** |
| **Cancer** | **4,678 (3,592, 5,799)** | **4.8** | **2,319 (1,712, 2,913)** | **4.1** | **2,360 (1,815, 2,904)** | **5.9** |
| Pancreas (C25) | 1,064 (224, 1,919) | 1.1 | 602 (126, 1,078) | 1.1 | 469 (88, 830) | 1.2 |
| Colorectum (C18-20) | 1,829 (1,249, 2,428) | 1.9 | 1,050 (720, 1,387) | 1.9 | 780 (532, 1,021) | 1.9 |
| Breast (C50) | 206 (-63, 466) | 0.2 | 0 | 0.0 | 204 (-70, 482) | 0.5 |
| Endometrium (C54.1) | 577 (427, 723) | 0.6 | 0 | 0.0 | 579 (417, 726) | 1.4 |
| Gallbladder (C23) | 101 (68, 133) | 0.1 | 38 (25, 49) | 0.1 | 64 (42, 83) | 0.2 |
| Kidney (C64) | 906 (712, 1,087) | 0.9 | 630 (503, 757) | 1.1 | 274 (217, 329) | 0.7 |
| **Chronic renal failure (N18)** | **3,615 (1,879, 5,241)** | **3.7** | **2,012 (1,006, 2,942)** | **3.6** | **1,597 (830, 2,338)** | **4.0** |
| **Liver disease (K70, K74)** | **4,487 (2,999, 5,832)** | **4.6** | **3,012 (2,057, 3,954)** | **5.3** | **1,452 (975, 1,875)** | **3.6** |

| **Cause of death (ICD-10 Code) <sup>1</sup>** | **Total (mean, 95% UI) <sup>2</sup>** | **%** | **Males (mean, 95% UI) <sup>2</sup>** | **%** | **Females (mean, 95% UI) <sup>2</sup>** | **%** |
| --- | --- | --- | --- | --- | --- | --- |
| **Total deaths averted or delayed** | **137,261 (125,534, 148,719)** | **100.0** | **78,295 (71,267, 85,112)** | **100** | **58,910 (53,288, 64,506)** | **100** |
| *Total deaths prevented under 75* | *55,744 (50,930, 60,515)* | *40.6* | *38,039 (34,474, 41,525)* | *48.6* | *17,675 (16,033, 19,299)* | *30.0* |
| **Cardiovascular diseases** | **101,669 (90,812, 112,460)** | **74.1** | **57,921 (51,576, 63,926)** | **74.0** | **43,680 (38,534, 48,855)** | **74.1** |
| Ischaemic heart disease (I20-25) | 44,689 (39,909, 49,671) | 32.6 | 29,979 (26,500, 33,412) | 38.3 | 14,711 (11,924, 17,444) | 25.0 |
| Cerebrovascular disease (I60-69) | 15,520 (12,330, 18,586) | 11.3 | 7,113 (5,611, 8,511) | 9.1 | 8,369 (6,673, 10,117) | 14.2 |
| Heart failure (I50) | 16,561 (11,751, 21,068) | 12.1 | 8,064 (5,659, 10,297) | 10.3 | 8,501 (6,038, 10,776) | 14.4 |
| Aortic aneurysm (I71) | 182 (78, 307) | 0.1 | 119 (51, 197) | 0.2 | 62 (25, 103) | 0.1 |
| Pulmonary embolism (I26) | 82 (26, 165) | 0.1 | 44 (14, 90) | 0.1 | 37 (12, 75) | 0.1 |
| Rheumatic heart disease (I05-09) | 30 (8, 62) | 0.0 | 12 (4, 26) | 0.0 | 17 (5, 35) | 0.0 |
| Hypertensive disease (I10-15) | 24,632 (19,702, 28,835) | 17.9 | 12,559 (10,121, 14,880) | 16.0 | 12,061 (9,743, 14,144) | 20.5 |
| **Diabetes (E11, E14)** | **17,786 (14,435, 20,687)** | **13.0** | **10,440 (8,364, 12,249)** | **13.3** | **7,345 (5,990, 8,492)** | **12.5** |
| **Cancer** | **6,696 (5,056, 8,212)** | **4.9** | **3,199 (2,370, 4,024)** | **4.1** | **3,471 (2,635, 4,275)** | **5.9** |
| Pancreas (C25) | 1,530 (288, 2,707) | 1.1 | 820 (161, 1,473) | 1.0 | 696 (133, 1,239) | 1.2 |
| Colorectum (C18-20) | 2,620 (1,784, 3,442) | 1.9 | 1,462 (984, 1,908) | 1.9 | 1,166 (780, 1,533) | 2.0 |
| Breast (C50) | 263 (-137, 651) | 0.2 | 0 | 0.0 | 269 (-133, 668) | 0.5 |
| Endometrium (C54.1) | 850 (621, 1,068) | 0.6 | 0 | 0.0 | 850 (623, 1,061) | 1.4 |
| Gallbladder (C23) | 147 (100, 192) | 0.1 | 52 (34, 68) | 0.1 | 94 (64, 124) | 0.2 |
| Kidney (C64) | 1,272 (1,015, 1,528) | 0.9 | 869 (688, 1,041) | 1.1 | 406 (325, 485) | 0.7 |
| **Chronic renal failure (N18)** | **5,027 (2,534, 7,220)** | **3.7** | **2,730 (1,329, 3,890)** | **3.5** | **2,291 (1,159, 3,305)** | **3.9** |
| **Liver disease (K70, K74)** | **6,267 (4,172, 8,137)** | **4.6** | **4,133 (2,749, 5,420)** | **5.3** | **2,151 (1,413, 2,752)** | **3.7** |

<sup>1</sup>WHO, International Statistical Classification of Diseases and Related Health Problems, Tenth Revision.
<sup>2</sup>95% Uncertainty Interval (UI) are based on 10,000 iterations of Monte Carlo analysis built in PRIME.
Total deaths averted or delayed represent less than the sum of the individual diet related NCD mortality causes given that double
counting has been accounted for in PRIME during the modelling process. The same applies to the sum of CVDs and cancers.
Potential diet related NCD deaths that could be averted or delayed were estimated using the PRIME model[20]. Inputs for the model included, 1) population demographics; 2) mortality data associated with diet related NCDs, obtained from the publicly available Centers for Disease Control and Prevention (CDC) WONDER datasets, 2019[29, 30]; and 3) baseline and counterfactual dietary intakes.

Of the total diet related NCD deaths that could have been prevented or delayed, 73.6% were related to CVDs, followed by diabetes (13.4%), cancers (4.8%), liver disease (4.6%), and chronic renal failure (3.7%). Furthermore, 39.4% of potential deaths averted or delayed would be in individuals under 75 years old. (**Table 3**).

#### Scenario 2: based on a meta-analysis on the impact of nutrient warning FOPL

Under scenario 2 (S2) based on results from a meta-analysis of randomized controlled trials and quasi-experimental studies investigating the impact of nutrient warning FOPL[6], we projected a mean (SE) usual total energy intake of 2,020 (14) kcal per day. This resulted in a reduction of 57 calories per day compared to the observed (baseline) scenario, considering only the calorie contributions from changes observed in sugar and saturated fat. However, this reduction was not statistically significant. Additionally, we observed absolute mean dietary reductions of 259 mg/day sodium (−7.7%), 7.2 g/day sugars (−7.1%), and 4.5 g/day saturated fats (−16.3%). The reductions in sodium, sugar, and saturated fat intakes were statistically significant (**Table 2**). Stratified results by DRI age-sex group were also estimated and are presented in **Supplementary Table S2**.

Estimated dietary intakes changes could avert or delay 137,261 (95% UI 125,534, 148,719) diet related NCD deaths in the US. Overall, more lives would be saved in males than females. Approximately 57% of averted or delayed deaths were estimated in males (78,295 [95% UI 71,267, 85,112]) and 43% in females (58,910 [95% UI 53,288, 64,506] (**Table 3**).

Of the total diet related NCD deaths that could have been prevented or delayed, 74.1% were related to CVDs, followed by diabetes (13.0%), cancers (4.9%), liver disease (4.6%), and chronic renal failure (3.7%). Furthermore, 40.6% of potential deaths averted or delayed would be in individuals under 75 years old (**Table 3**).

#### Potential dietary impacts by DRI age-sex groups

When examining by DRI age-sex group, significant sodium reductions were observed in males ages 19-30 y (S2), 31-50 y (S1, S2), 51-70 y (S2), and 71+ y (S2); and in females ages 19-30 y (S2), 31-50 y (S2), and 51-70 y (S2). For total sugars, significant reductions were observed only in females ages 19-30 y (S1), 31-50 y (S1, S2), 51-70 y (S1, S2), and 71+ y (S1). For saturated fat, significant changes were observed for no age or sex group under S1 but for all age and sex groups under S2. (**Supplementary Table S2).**

Of the total number of deaths that could be prevented or delayed in both scenarios, between 89% and 91% would be attributable to changes in energy intakes (changes in obesity status), between 9% and 11% would be attributable to changes in sodium intakes, and between 0.3% and 1.7% would be attributable to changes in saturated fat intakes.

### Sensitivity analysis

The mean (SE) usual added sugars intake per day among adults in the US ages 19 years and older was 66.8 (1.73) g/day (**Supplementary Table S2**). Applying potential intervention effects to added sugars and not to total sugars, as the main analysis, resulted in absolute mean added sugar reductions of 6.7 and 4.8 g/day under S1 and S2, respectively. The reduction in added sugar intake was only significant under S1. Estimated health gains were lower than the main analysis under both scenarios tested. The number of deaths that could have been prevented or delayed decreased by 31% under S1 and by 14% under S2 (**Supplementary Table S3**).

## Discussion

To the best of our knowledge, this is the first study to model potential dietary and health impacts of implementing a nutrient-specific FOPL policy in the US. This study estimated the potential dietary and NCD health impact among US adults in response to mandatory nutrient-specific FOPL regulations. Our results indicate that a FOPL policy in the US could potentially avert or delay more than 137,000 diet related NCD deaths among US adults, largely attributable to reductions in energy intake. This potential health impact is largely attributable to CVDs and type-2 diabetes. Our results demonstrate the potential impact that even modest changes in dietary intake may have on population health. If the US adult population responded to a FOPL policy similarly to populations in other countries[6, 9] we would expect a small but significant reduction in population-level sodium intake of up to 259 mg/day and a reduction in total sugar intake of 10 g/day.

Previous modeling studies in the US have estimated the potential impact of implementing a sugar-swetened beverage (SSB) health warning policy. It was estimated that this policy would reduce total energy intake by 31.2 kcal/day among US adults[35]. Our estimates showed a potential calorie reduction of between 38 and 57 kcal/day derived from reductions in sugar and saturated fat intake as a result of implementing a mandatory nutrient-specific FOPL that would appear on both foods and beverages, including SSB. This shows that our estimated intervention effects may be more conservative than previous estimates, given that we applied these reductions to all food products and not just SSBs, and may underestimate potential dietary and health gains from this policy[35]. Another study tested the same counterfactual scenarios in the Canadian context, finding a potential calorie reduction of between 43 and 59 kcal/day, but as expected, a smaller number of diet-realted NCD deaths than our findings (8,907 vs 137,261), given Canada’s smaller population size and lower burden of NCDs compared to the US[36]. Moreover, a study in France modeled the potential impact on NCD mortality of different FOPL systems that did not include a nutrient-specific ‘high in’ type FOPL. The study found that a significant number of NCD-related deaths could be prevented or delayed by implementing the Nutri-score (7,680) and the HSR system (6,265). However, the study assumed that these policies were mandatory, which is not the case at the moment, and there are no country-specific evaluations supporting this assumption[37].

The estimated reduction in nutrients-of-concern could result from both consumers making healthier food choices and initial food reformulation by the food industry. Changes in consumer behavior might involve selecting similar products with lower levels of nutrients-of-concern (i.e., fewer or no nutrient-specific FOPL), discontinuing consumption of certain products that are high in nutrients-of-concern, or increasing consumption of healthier products. For instance, under S1 based on initial evaluations of Chile’s FOPL policy, overall reductions applied in our policy scenario were a result of estimated reductions in consumption of ‘high in’ products and increased consumption of not ‘high in’ products, according to Taillie, et al[9].

Furthermore, industry-driven initial food reformulation has been observed as a response to mandatory nutrient-specific ‘high in’ FOPL, with manufacturers modifying their products to avoid displaying ‘high in’ FOPL. In Chile, a significant decrease was observed in the proportion of products displaying any ‘high in’ label, with the most common reductions seen in the proportion of products displaying a ‘high in’ FOPL for sodium and sugar. For instance, the proportion of ‘high in’ sodium products decreased most frequently in food categories such as savory spreads, cheeses, ready-to-eat meals, soups, and sausages, dropping in some cases from 74% to 27%. Similarly, ‘high in’ sugar products saw reductions most frequently in categories such as beverages, milk and milk-based drinks, breakfast cereals, sweet baked products, and sweet and savory spreads, dropping in some cases from 80% to 60%[11]. Some of these food categories are also top contributors to sodium and added sugar intake in the US[38, 39], which highlights the potential for industry-driven food reformulation as a result of adopting and implementing a mandatory nutrient-specific ‘high in’ FOPL in the US. However, robust and independent monitoring efforts are warranted to monitor changes in the US packaged food supply and any unintended consequences from food reformulation. For example, in Chile, the use of non-nutritive sweeteners (NNS) in the food and beverage supply increased from 37.9% to 43.6% following the law’s implementation[40]. Lessons from the Chilean experience informed other nutrient-specific ‘excess’ FOPL regulations in countries such as Mexico and Argentina that also require precautionary labels for products containing NNS[41, 42].

The nature of the FOPL policy (mandatory vs. voluntary), FOPL design, and underlying NPM are of great relevance to consider by policymakers adopting and implementing FOPL. In the US, where no such policy exists, it is important that future policy designs are informed by prior research and country experiences on what is most likely to optimize consumers’ use and understanding of the labels, promote healthier food choices, and consequently improve the population’s health. For instance, mandatory nutrient-specific ‘high in’ or ‘excess’ FOPL, like the one implemented in Chile and several other countries [43], has been shown to effectively change consumer purchasing behaviors [8, 9], encourage industry-driven food reformulation [10, 11], and improve diets [12]. Additionally, initial evaluations from countries that recently implemented similar policies demonstrate consumers’ use and comprehension of nutrient-specific ‘high in’ FOPL[44–48] and a reduction in the proportion of food and beverage proucts displaying any ‘high in’ FOPL after policy implementation[49]. In contrast, recent evaluations of the voluntary Health Star Rating (HSR) system, implemented in Australia and New Zealand in 2014, showed no impact on consumer purchasing behavior[50].Moreover, only 25% of foods were labeled with the HSR label in 2019, with HSR-labeled products more likely to display a 4.0-5.0 rating to highligth healthier alternatives, suggesting that companies are selectively applying HSR labels as a marketing tool as opposed to a public health policy[50]. This demonstrates the greater effectiveness of mandatory approaches compared to voluntary ones.

When interpreting our results, it is important to consider both the strengths and limitations. A strength of our study is the use of data from the nationally representative NHANES (2017-2020) survey to estimate our baseline and counterfactual scenarios. Although 24HR are subject to misreporting issues, the US Department of Agriculture Automated Multiple Pass Method was used to reduce misreporting bias[51]. Additionally, we employed both 24HR days and the NCI method to evaluate usual calorie and nutrient intakes for US adults, overall and by DRI age-sex group, adjusting for various relevant covariates. This approach enabled the use of disaggregated data by DRI age-sex group into PRIME. The counterfactual scenarios in this study were modeled using the latest evidence on changes in food and beverage purchases when a nutrient-specific ‘high in’ FOPL is present. Due to the limited evidence on how FOPL policies affect dietary intakes, it was assumed that the observed changes in calories and critical nutrients in food and beverage purchases with a nutrient-specific FOPL would translate to dietary intakes. This assumption is supported by research comparing food purchase data with 24HR data, which suggests that recorded food purchases can reasonably estimate overall diet quality[52].

We modeled the potential dietary and health impact of implementing nutrient-specific ‘high in’ FOPL using evidence from Chile, which, to our knowledge, is the only country that has evaluated changes in food and beverage purchases before and after implementing such policy. The changes observed in Chile may not directly apply to the US context due to several differences, such as dietary patterns, cultural food norms, food environments, and differences in labeling regulations between the two countries, but the impact and direction of these various differences is unclear. However, both Chilean and US populations share similar diet related risk factors and burden of obesity. For example, both countries have among the highest per capita retail sales of ultra-processed products [53], high sodium intakes [54, 55], and a high prevalence of obesity [56].

Furthermore, it is important to note that the comprehensive Chilean law not only includes a mandatory ‘high in’ FOPL but also restricts sales and promotions in schools and marketing to children for foods that exceed established thresholds for calorie density and targeted nutrients, policies that have not yet been considered in the US. Consequently, initial evaluations of the Chilean law likely reflect the impact of these additional components. Policy scenarios that consider trends in consumer purchase behavior following the implementation of mandatory nutrient-specific FOPL should be further investigated as more evidence on the long-term impacts of this policy becomes available in other countries such as Mexico, Argentina, and Colombia. Specifically, it is important to determine whether the effects on consumer food choices level off or reverse over time and, if so, at what point this occurs. Additionally, the NPM adopted for FOPL regulations in Chile might be different to what will be proposed by the FDA, likely resulting in fewer products displaying FOPL nutrient disclosures in the US compared to Chile. Future research should inform how the US packaged food supply would perform under the proposed and other evidence-based, well-established NPMs.

Lastly, as other cross-sectional NCD scenario models, PRIME does not account for the effect of time lag between the exposure and disease outcome, making it unclear how long it would take for the estimated health gains to manifest following a change in risk factor exposure.

Nonetheless, the model has been used extensively as it enables researchers to estimate the population-level health impact of various NCD policy scenarios, providing crucial evidence for informing the policymaking process and prioritizing resources as necessary. The strengths and limitations of PRIME have been discussed in detail before[20, 57]. Future studies should build on this work to estimate the cost-effectiveness of this policy in the US context and examine potential differential impacts across diverse subgroups.

## Conclusion

Findings suggest that implementing mandatory nutrient-specific FOPL in the US could significantly reduce sodium and total sugar intakes among US adults. This change could potentially avert or delay up to 137,261 diet related NCD deaths in the US, primarily from CVDs. Our results can inform current food policy developments in the US regarding adoption and implementation of FOPL regulations.

## Data Availability

All relevant data are within the manuscript and its Supporting Information files.

## Authors’ contributions

Conceptualization, NF, and ML; Data Curation, NF, and DZ; Formal Analysis, NF and DZ; Investigation, NF, and DZ; Funding Acquisition, ML, and NF; Methodology, NF, MA, and DZ; Writing – Original Draft Preparation, NF and DZ. All authors critically reviewed and approved the final manuscript. EG, and AM are staff members of the Center for Science in the Public Interest. Authors hold sole responsibility for the views expressed in the manuscript, which may not necessarily reflect the opinion or policy of their respective organizations.

## Acknowledgments

The authors would like to thank Professor Peter Scarborough, University of Oxford, for allowing us to use the PRIME model and discussing its application.

## References

1. Afshin A, Sur PJ, Fay KA, Cornaby L, Ferrara G, Salama JS, et al. Health effects of dietary risks in 195 countries, 1990&#x2013;2017: a systematic analysis for the Global Burden of Disease Study 2017. The Lancet. doi: 10.1016/S0140-6736(19)30041-8.

2. Baldridge AS, Huffman MD, Taylor F, Xavier D, Bright B, Van Horn LV, et al. The healthfulness of the US packaged food and beverage supply: A cross-sectional study. Nutrients. 2019;11(8):1704.

3. Roberto CA, Ng SW, Ganderats-Fuentes M, Hammond D, Barquera S, Jauregui A, Taillie LS. The Influence of Front-of-Package Nutrition Labeling on Consumer Behavior and Product Reformulation. Annu Rev Nutr. 2021;41(Volume 41, 2021):529–50. doi: 10.1146/annurev-nutr-111120-094932.

4. Global Food Research Program. Front-of-package labeling policies around the world 2024 [cited 2024 May 22]. Available from: https://www.globalfoodresearchprogram.org/wp-content/uploads/2022/07/GFRP-UNC_FOPL_maps_2024_03.pdf.

5. Crosbie E, Gomes FS, Olvera J, Rincón-Gallardo Patiño S, Hoeper S, Carriedo A. A policy study on front–of–pack nutrition labeling in the Americas: Emerging developments and outcomes. The Lancet Regional Health - Americas. 2022:100400. doi: 10.1016/j.lana.2022.100400.

6. Song J, Brown MK, Tan M, MacGregor GA, Webster J, Campbell NR, et al. Impact of color-coded and warning nutrition labelling schemes: A systematic review and network meta-analysis. PLoS Medicine. 2021;18(10):e1003765.

7. Croker H, Packer J, Russell SJ, Stansfield C, Viner R. Front of pack nutritional labelling schemes: a systematic review and meta-analysis of recent evidence relating to objectively measured consumption and purchasing. Journal of Human Nutrition and Dietetics. 2020.

8. Taillie LS, Reyes M, Colchero MA, Popkin B, Corvalán C. An evaluation of Chile’s Law of Food Labeling and Advertising on sugar-sweetened beverage purchases from 2015 to 2017: A before-and-after study. PLoS medicine. 2020;17(2):e1003015.

9. Taillie LS, Bercholz M, Popkin B, Reyes M, Colchero MA, Corvalán C. Changes in food purchases after the Chilean policies on food labelling, marketing, and sales in schools: a before and after study. The Lancet Planetary Health. 2021;5(8):e526–e33.

10. Quintiliano Scarpelli D, Pinheiro Fernandes AC, Rodriguez Osiac L, Pizarro Quevedo T. Changes in nutrient declaration after the food labeling and advertising law in Chile: a longitudinal approach. Nutrients. 2020;12(8):2371.

11. Reyes M, Smith Taillie L, Popkin B, Kanter R, Vandevijvere S, Corvalán C. Changes in the amount of nutrient of packaged foods and beverages after the initial implementation of the Chilean Law of Food Labelling and Advertising: A nonexperimental prospective study. PLOS Medicine. 2020;17(7):e1003220. doi: 10.1371/journal.pmed.1003220.

12. Fretes G, Corvalán C, Reyes M, Taillie LS, Economos CD, Wilson NLW, Cash SB. Changes in children’s and adolescents’ dietary intake after the implementation of Chile’s law of food labeling, advertising and sales in schools: a longitudinal study. International Journal of Behavioral Nutrition and Physical Activity. 2023;20(1):40. doi: 10.1186/s12966-023-01445-x.

13. Taillie LS, Bercholz M, Popkin B, Rebolledo N, Reyes M, Corvalán C. Decreases in purchases of energy, sodium, sugar, and saturated fat 3 years after implementation of the Chilean food labeling and marketing law: An interrupted time series analysis. PLOS Medicine. 2024;21(9):e1004463. doi: 10.1371/journal.pmed.1004463.

14. Ministerio de Salud de Chile. Ley 20606 Sobre composicion nutricional de los alimentos y su publicidad 2012 [cited 2024 September 28]. Available from: https://www.bcn.cl/leychile/navegar?i=1041570&f=2012-07-06&p=.

15. Correa T, Reyes M, Taillie LS, Corvalán C, Dillman Carpentier FR. Food advertising on television before and after a national unhealthy food marketing regulation in chile, 2016–2017. Am J Public Health. 2020;110(7):1054–9.

16. Martin C, Turcotte M, Cauchon J, Lachance A, Pomerleau S, Provencher V, Labonté M-È. Systematic review of nutrient profile models developed for nutrition-related policies and regulations aimed at noncommunicable disease prevention–An update. Advances in Nutrition. 2023.

17. Global Health Advocacy Incubator. Nutrient Profile Models: A valuable tool for developing healthy food policies 2024 [cited 2024 May 23]. Available from: https://assets.advocacyincubator.org/uploads/2024/NPM_Position_Paper.pdf.

18. The White House. Biden-Harris Administration National Strategy on Hunger, Nutrition, and Health 2022 [cited 2024 June 20]. Available from: https://www.whitehouse.gov/wp-content/uploads/2022/09/White-House-National-Strategy-on-Hunger-Nutrition-and-Health-FINAL.pdf

19. US Food and Drug Administration. Front-of-Package Nutrition Labeling 2024 [cited 2024 May 23]. Available from: https://www.fda.gov/food/food-labeling-nutrition/front-package-nutrition-labeling.

20. Scarborough P, Harrington RA, Mizdrak A, Zhou LM, Doherty A. The preventable risk integrated ModEl and its use to estimate the health impact of public health policy scenarios. Scientifica. 2014;2014.

21. Stierman B, Afful J, Carroll MD, Chen T-C, Davy O, Fink S, et al. National Health and Nutrition Examination Survey 2017–March 2020 prepandemic data files development of files and prevalence estimates for selected health outcomes. 2021.

22. Willett W. Issues in analysis and presentation of dietary data. Nutritional epidemiology. 1998:321–46.

23. U.S. Department of Agriculture and U.S. Department of Health and Human Services. Dietary Guidelines for Americans, 2020-2025 2020. 9th Edition:[Available from: https://www.dietaryguidelines.gov/sites/default/files/2021-03/Dietary_Guidelines_for_Americans-2020-2025.pdf.

24. Office of Information and Regulatory Affairs. (CFSAN) Front-of-Package Nutrition Labeling Focus Groups 2. Appendix A - FOP Focus Groups 2 Schemes and Product MockUps 8-23-2023 2024 [cited 2024 May 23]. Available from: https://www.reginfo.gov/public/do/PRAViewIC?ref_nbr=202008-0910-021&icID=262002.

25. National Academies of Sciences Engineering and Medicine. Dietary Reference Intakes Collection 2022 [cited 2022 October 12]. Available from: https://nap.nationalacademies.org/collection/57/dietary-reference-intakes.

26. Cifelli CJ, Houchins JA, Demmer E, Fulgoni VL. Increasing plant based foods or dairy foods differentially affects nutrient intakes: Dietary scenarios using NHANES 2007–2010. Nutrients. 2016;8(7):422.

27. Quann EE, Fulgoni VL, Auestad N. Consuming the daily recommended amounts of dairy products would reduce the prevalence of inadequate micronutrient intakes in the United States: diet modeling study based on NHANES 2007–2010. Nutrition journal. 2015;14(1):1–11.

28. World Health Organization - Regional Office for Europe. NCDprime - Modelling the impact of national policies on noncommunicable disease (NCD) mortality using PRIME: a policy scenario modelling tool (2019) 2019 [cited 2023 October 11]. Available from: https://www.who.int/europe/publications/i/item/WHO-EURO-2019-3652-43411-60952.

29. Centers for Disease Control and Prevention CDC WONDER. Bridged-Race Population Estimates 1990-2019 Request 2019 [cited 2024 March 9]. Available from: https://wonder.cdc.gov/bridged-race-v2019.html.

30. Centers for Disease Control and Prevention CDC WONDER. Underlying Cause of Death, 1999-2020 Request. Deaths occurring through 2020 2019 [cited 2024 March 9]. Available from: https://wonder.cdc.gov/ucd-icd10.html.

31. World Health Organization. International Statistical Classification of Diseases and Related Health Problems 10th Revision 2016 [cited 2021 January 4]. Available from: https://icd.who.int/browse10/2016/en#/I20-I25.

32. National Cancer Institute. Usual Dietary Intakes: SAS Macros for the NCI Method 2018 [cited 2024 September 20]. Available from: https://epi.grants.cancer.gov/diet/usualintakes/macros.html.

33. Herrick KA, Rossen LM, Parsons R, Dodd KW. Estimating usual dietary intake from National Health and Nutrition Examination Survey data using the National Cancer Institute method. 2018.

34. World Health Organization. Obesity and overweight, Key Facts 2024 [cited 2024 June 21]. Available from: https://www.who.int/news-room/fact-sheets/detail/obesity-and-overweight.

35. Grummon AH, Smith NR, Golden SD, Frerichs L, Taillie LS, Brewer NT. Health warnings on sugar-sweetened beverages: simulation of impacts on diet and obesity among US adults. American journal of preventive medicine. 2019;57(6):765–74.

36. Flexner N, Ng AP, Ahmed M, Khandpur N, Acton RB, Lee JJ, L’Abbe MR. Estimating the dietary and health impact of implementing front-of-pack nutrition labeling in Canada: A macrosimulation modeling study. Frontiers in Nutrition. 2023;10. doi: 10.3389/fnut.2023.1098231.

37. Egnell M, Crosetto P, d’Almeida T, Kesse-Guyot E, Touvier M, Ruffieux B, et al. Modelling the impact of different front-of-package nutrition labels on mortality from non-communicable chronic disease. Int J Behav Nutr Phys Act. 2019;16(1):56. Epub 2019/07/17. doi: 10.1186/s12966-019-0817-2. PubMed PMID: 31307496; PubMed Central PMCID: PMCPMC6631735.

38. Lee SH, Zhao L, Park S, Moore LV, Hamner HC, Galuska DA, Blanck HM. High added sugars intake among US adults: Characteristics, eating occasions, and top sources, 2015–2018. Nutrients. 2023;15(2):265.

39. Ahmed M, Ng A, Christoforou A, Mulligan C, L’Abbé MR. Top sodium food sources in the American diet—using National Health and Nutrition Examination Survey. Nutrients. 2023;15(4):831.

40. Zancheta Ricardo C, Corvalán C, Taillie L, Quitral V, Reyes M. Changes in the use of nonnutritive sweeteners in the Chilean food and beverage supply after the implementation of the Food Labeling and Advertising Law. Frontiers in nutrition. 2021:906.

41. White M, Barquera S. Mexico Adopts Food Warning Labels, Why Now? Health Systems & Reform. 2020;6(1):e1752063.

42. República Argentina. Ley de etiquetado frontal. Promoción de la alimentación saludable Ley 27.642. 2022 [cited 2022 October 12]. Available from: https://www.argentina.gob.ar/justicia/derechofacil/leysimple/salud/ley-de-etiquetado-frontal#:~:text=Est%C3%A1%20prohibida%20la%20publicidad%2C%20promoci%C3%B3n,a%20ni%C3%B1os%2C%20ni%C3%B1as%20y%20adolescentes.

43. Global Health Advocacy Incubator. FOPWL regulations around the globe 2023 [cited 2023 October 2]. Available from: https://dfweawn6ylvgz.cloudfront.net/uploads/2023/FOPWL_Regulations.pdf.

44. Contreras-Manzano A, White CM, Nieto C, Quevedo KL, Vargas-Meza J, Hammond D, et al. Self-reported decreases in the purchases of selected unhealthy foods resulting from the implementation of warning labels in Mexican youth and adult population. International Journal of Behavioral Nutrition and Physical Activity. 2024;21(1):64.

45. Shahrabani S. The impact of Israel’s Front-of-Package labeling reform on consumers’ behavior and intentions to change dietary habits. Israel Journal of Health Policy Research. 2021;10(1):1–11.

46. Ares G, Antúnez L, Curutchet MR, Galicia L, Moratorio X, Giménez A, Bove I. Immediate effects of the implementation of nutritional warnings in Uruguay: awareness, self-reported use and increased understanding. Public Health Nutrition. 2021;24(2):364–75.

47. Machín L, Alcaire F, Antúnez L, Giménez A, Curutchet MR, Ares G. Use of nutritional warning labels at the point of purchase: An exploratory study using self-reported measures and eye-tracking. Appetite. 2023:106634.

48. Batis C, Aburto TC, Pedraza LS, Angulo E, Hernández Z, Jáuregui A, et al. Self-reported reactions to the front-of-package warning labelling in Mexico among parents of school-aged children. medRxiv. 2023:2023.07. 26.23293213.

49. Saavedra-Garcia L, Meza-Hernández M, Diez-Canseco F, Taillie LS. Reformulation of Top-Selling Processed and Ultra-Processed Foods and Beverages in the Peruvian Food Supply after Front-of-Package Warning Label Policy. International Journal of Environmental Research and Public Health. 2022;20(1). doi: 10.3390/ijerph20010424.

50. Bablani L, Mhurchu CN, Neal B, Skeels CL, Staub KE, Blakely T. Effect of voluntary Health Star Rating labels on healthier food purchasing in New Zealand: longitudinal evidence using representative household purchase data. BMJ Nutrition, Prevention & Health. 2022;5(2).

51. Moshfegh AJ, Rhodes DG, Baer DJ, Murayi T, Clemens JC, Rumpler WV, et al. The US Department of Agriculture Automated Multiple-Pass Method reduces bias in the collection of energy intakes. The American journal of clinical nutrition. 2008;88(2):324–32.

52. Appelhans BM, French SA, Tangney CC, Powell LM, Wang Y. To what extent do food purchases reflect shoppers’ diet quality and nutrient intake? International Journal of Behavioral Nutrition and Physical Activity. 2017;14(1):1–10.

53. Pan American health Organization. Ultra-processed food and drink products in Latin America: Trends, impact on obesity, policy implications 2015 [cited 2023 October 5]. Available from: https://iris.paho.org/bitstream/handle/10665.2/7699/9789275118641_eng.pdf?sequence=5&isAllowed=y.

54. Gobierno de Chile - Ministerio de Salud. Encuesta Nacional de Salud 2016-2017. Segunda entrega de resultados. 2018 [cited 2024 May 25]. Available from: https://redsalud.ssmso.cl/wp-content/uploads/2018/02/2-Resultados-ENS_MINSAL_31_01_2018-ilovepdf-compressed.pdf.

55. Brouillard AM, Kraja AT, Rich MW. Trends in dietary sodium intake in the United States and the impact of USDA guidelines: NHANES 1999-2016. The American journal of medicine. 2019;132(10):1199–206. e5.

56. World Obesity. Global Obesity Observatory. WHO Americas region 2022 [cited 2024 May 25]. Available from: https://data.worldobesity.org/region/who-americas-region-3/#data_prevalence.

57. Dötsch-Klerk M, Bruins MJ, Detzel P, Martikainen J, Nergiz-Unal R, Roodenburg AJ, Pekcan AG. Modelling health and economic impact of nutrition interventions: a systematic review. European Journal of Clinical Nutrition. 2022:1–14.

